# Reinfection Rates among Patients who Previously Tested Positive for COVID-19: a Retrospective Cohort Study

**DOI:** 10.1101/2021.02.14.21251715

**Authors:** Megan M. Sheehan, Anita J. Reddy, Michael B. Rothberg

## Abstract

**Objectives:** To evaluate reinfection rates and protective effectiveness of prior disease among patients with coronavirus disease 2019 (COVID-19) infection in the United States.

**Design:** Retrospective cohort study

**Setting:** One multi-hospital health system in Ohio and Florida

**Participants:** All 150,325 patients who were tested for COVID-19 infection via PCR from March 12, 2020 to August 30, 2020. Testing performed up to January 7, 2021 in these patients was included for analysis. Healthcare workers were excluded.

**Main outcome measures:** The main outcome was reinfection, defined as infection ≥ 90 days after initial testing. Secondary outcomes were symptomatic infection and protective effectiveness of prior infection.

**Results:** Of 150,325 patients tested for COVID-19 prior to August 30, 8,845 (5.9%) tested positive and 141,480 (94.1%) tested negative. 974 (11%) of the positive patients were retested after 90 days, and 56 had possible reinfection. Of those, 26 (46.4 %) were symptomatic. Of those with initial negative testing, 4,163 (12.9%) were subsequently positive and 2,460 of those (59.1%) were symptomatic. Protective effectiveness of prior infection was 78.5% (95% confidence interval 72.0 to 83.5), and against symptomatic infection was 83.1% (95% confidence interval 75.1 to 88.5). Protective effectiveness increased over time.

**Conclusions:** Prior infection in patients with COVID-19 was highly protective against reinfection and symptomatic disease. Protective effectiveness increased over time, suggesting that viral shedding or ongoing immune response may persist beyond 90 days and may not represent true reinfection. As vaccine supply is a limited resource around the world, patients with known history of COVID-19 could delay early vaccination to allow for the most vulnerable to access the vaccine and slow transmission.

## Introduction

SARS-CoV-2 has infected >27 million Americans, nearly 4 million in the United Kingdom, and >107 million individuals worldwide as of February 12, 2021. This number is likely underestimated due to limited testing and lack of surveillance for asymptomatic infections.^1^ The protection afforded by SARS-CoV-2 infection remains unknown. There are several reports of reinfection with phylogenetically distinct variants of SARS-CoV-2, including in the United States, but these are rare.^2–4^ Studies of patients infected during the SARS pandemic of 2003 suggest that antibody response from infection persists over 2 years.^5^ In contrast, infection with common seasonal strains of human coronavirus does not confer lasting protection against reinfection, although reinfection within 6 months is uncommon.^6^

There are now several vaccines that have been licensed in various countries. Vaccine efficacy ranges from 62% to 95%.^7,8^ Due to shortages in vaccine supply, almost all countries have created prioritization schemes to ensure that those at highest risk from the virus (i.e. elderly patients and those with co-morbidities) and front-line healthcare workers receive the vaccine first. But demand remains high and additional strategies, such as administering only one dose of the two-dose regimen, have been proposed.^9,10^

Given the widespread nature of the pandemic and the lack of available testing, it is likely that a substantial portion of the population has already been infected with COVID-19. If such infection offered long-term protection, then vaccination of previously infected persons could be delayed until there is sufficient supply to vaccinate the entire population. Unfortunately, information on the long-term immunity conferred by infection with SARS-CoV-2 is scant. Current Centers for Disease Control (CDC) guidelines make no exceptions for persons with prior history of SARS-CoV-2 infection.^11^ However, if infection provides substantial long-lasting immunity, it may be appropriate to reconsider this recommendation. In order to help answer this question, we examined reinfection rates among a large number of patients with documented COVID-19 infection.

## Methods

### Subjects and outcomes

Patients tested for COVID-19 infection via PCR at one health system in Ohio and Florida from March 12, 2020 to January 7, 2021 were included. Health system employees were excluded due to privacy concerns. Initial infection status was based on tests performed prior to August 30, 2020. The primary outcome was a positive PCR test following the initial test. Per CDC definition, retesting and reinfection is defined as occurring ≥ 90 days after initial testing.^12^ Therefore, for patients who initially tested positive, we considered any positive test >90 days after the initial infection to be a reinfection, and ignored any repeat positive test within 90 days. To avoid bias, patients with baseline negative status who tested positive within 90 days of their initial test were excluded. Reinfections were reviewed to determine symptoms and severity. Each test had an indication and presence of symptoms recorded by the ordering provider, which was used to determine if the patient was symptomatic.

### Data Analysis

For each group of patients, we determined the reinfection rate, using the total number of patients in that group as the denominator. Infection rates were determined for distinct periods following the initial test: 4-5 months, 6-7 months and ≥ 8 months. Protective effectiveness of prior infection was calculated as one minus the ratio of infection rate for positive patients divided by the infection rate for negative patients. We determined the effectiveness in each period and overall. We then repeated the analysis including only symptomatic infections in the numerator. Analyses were conducted using R v.4.02 (R Core Team, Vienna). This work was approved by Cleveland Clinic’s Institutional Review Board (IRB 20-1328).

## Results

During the study period, 506,583 tests were collected from 333,606 individuals (average age 51.7 ± 22.2 years, 54.7% female), with a 9.8% overall positivity rate. Median tests per patient was 2 (IQR 1, 3), and 150,325 (45.1%) patients had tests performed before August 30. Of those, 8,845 (5.9%) individuals tested positive and 141,480 (94.1%) tested negative (Table). After at least 90 days, 974 (11%) of the positive patients were retested and 57 (5.9%) were reviewed for possible reinfection. One patient had an immediate negative test and was excluded due to a presumed false positive test.

**Table.** Characteristics of patients with initial positive tests compared to those with initial negative tests.

|  | **Initial Positive** | **Initial Negative** |
| --- | --- | --- |
| Number (patients) | 8845 | 141 480 |
| Age ±SD | 52.3 ± 21.8 | 54.8 ± 21.4 |
| Sex female (%) | 4605 (52.1) | 78 202 (55.1) |
| Number who were retested (%) | 974 (11) | 32 208 (22.8) |
| Age ±SD | 56.5 ± 20.2 | 57.1 ± 20.3 |
| Sex female (%) | 547 (56.2) | 18 034 (56) |
| Retest positive, any (%) | 56 (0.6) | 4163 (2.9) |
| Time to positive retest ±SD (days) | 131.4 ± 40.4 | 168.7 ± 46.4 |
| Retest positive, symptomatic (%) | 26 (0.3) | 2460 (1.7) |
|  | **Any infection** | **Symptomatic infection** |
| Effectiveness* | 78.5% | 83.1% |
| 90-150 days | 58.4% | 69.8% |
| 151-210 days | 91.3% | 91.8% |
| After 210 days | 92.7% | 91.1% |

\*Effectiveness = 1-((56/8845)/(4163/141480))

Of the 56 reinfections, 26 were symptomatic—shortness of breath was the most common symptom, and interestingly, no patient lost the sense of smell (Figure 1). Seventeen symptomatic patients were hospitalized within 30 days of the positive test, 5 with symptoms considered possibly related to COVID-19. Of those 5, none required intensive care or needed mechanical ventilation. Many reinfections occurred close to 90 days after initial infection, and average time to reinfection was 131.4 ± 40.4 days (range 90.2 – 269.0 days) (Figure 2).

**Figure 1.**
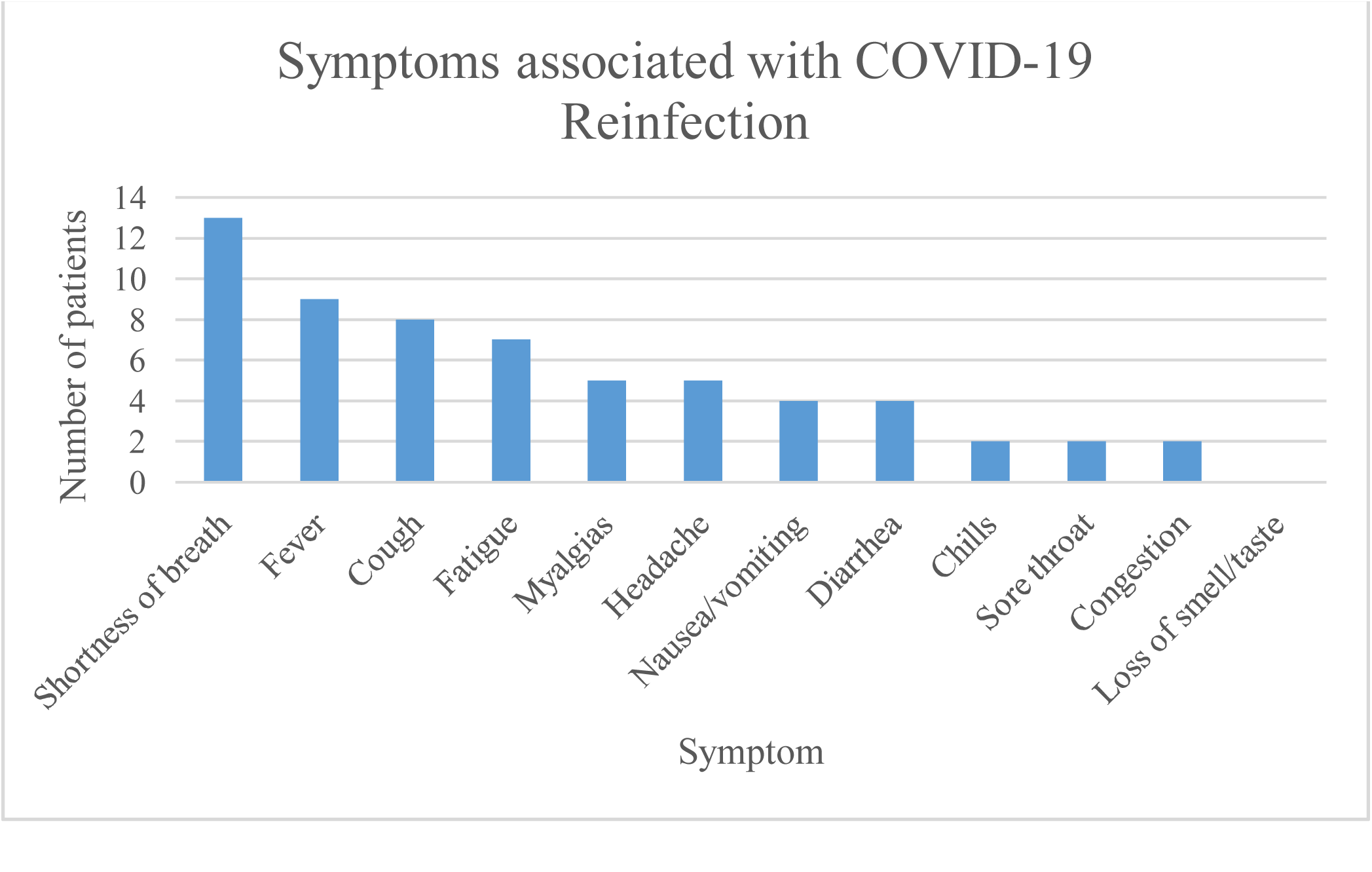
Symptoms of 26 patients with reinfection.

**Figure 2.**
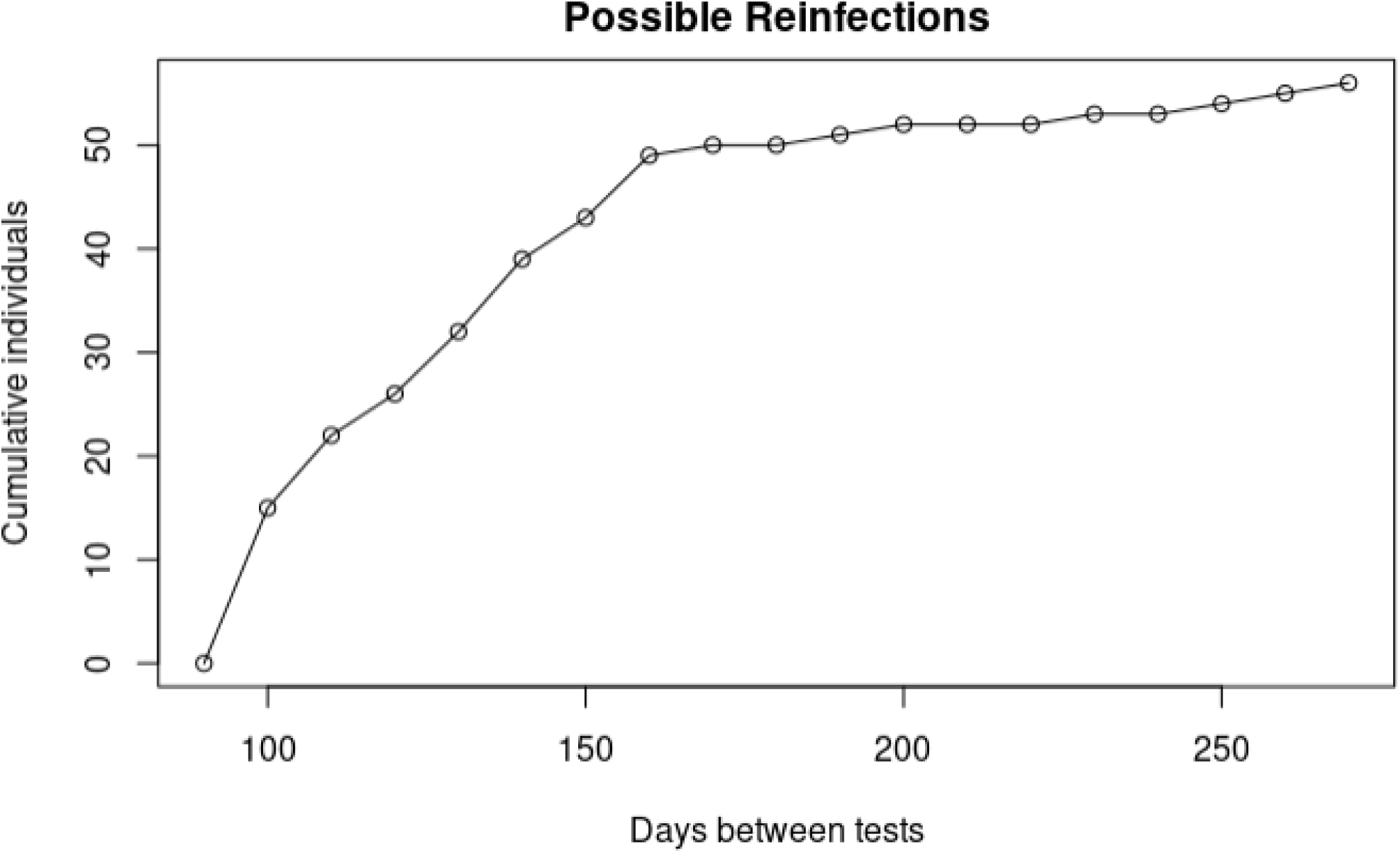
Time to reinfection for 56 patients.

Of those with negative initial tests, 22.8% (32,208/141,480) were retested and 4,163 (12.9%) were positive; 1703 (40.9%) positive tests were performed for pre-procedural screening or had an asymptomatic indication. The protective effectiveness of prior infection was 78.5% (95% CI 72.0, 83.5). Effectiveness against symptomatic infection was 83.1% (95% CI 75.1, 88.5). Risk of reinfection was greatest just after 90 days and declined thereafter (Figure 3). Consequently, effectiveness was lowest in months 4-5 and increased for up to 8 months after infection.

**Figure 3.**
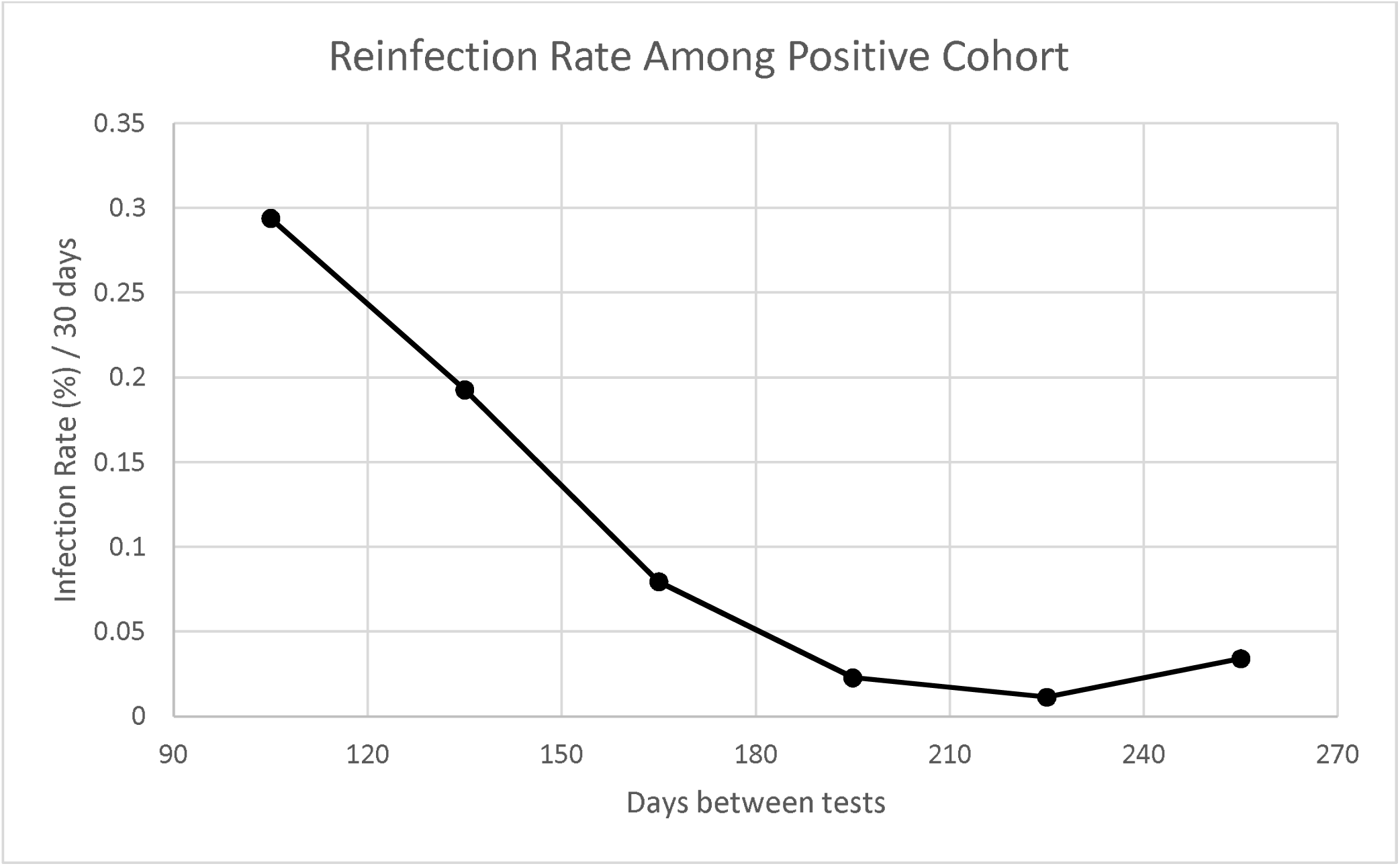
Reinfection rate over time for positive cohort.

## Discussion

In this retrospective cohort, patients who initially tested positive for COVID-19 were less likely to be subsequently tested or test positive than those who initially tested negative during the same time period. Most reinfected patients were asymptomatic. Protective effectiveness of prior infection against symptomatic disease was 83%, and even including asymptomatic cases, protective effectiveness was 78%. Few patients were hospitalized following reinfection, and none with COVID-related symptoms required intensive care, suggesting a high level of protection against severe disease. Six months after infection, protection against symptomatic disease exceeded 90%.

Others have estimated similar protectiveness of prior infection. Among healthcare workers in the UK, the presence of antibodies was associated with 91% reduced risk of symptomatic reinfection in the following 6 months,^13^ and in the Moderna vaccine study, previous infection afforded 76% protection in the placebo arm, although the numbers were exceedingly small (only one case of reinfection) and the confidence intervals were exceptionally wide.^8^

Our measure of reinfection may have overestimated the actual reinfection rate. Because some patients may continue to shed virus for many months, it can be difficult to differentiate between reinfection and persistent shedding. One cohort study found that 5.3% of participants were still positive at 90 days, which is substantially higher than what we observed, though most patients in our study were not retested.^14^ Patients with symptoms may be more likely to represent true reinfection, but the symptoms of COVID-19 that we observed were generally non-specific, and could have represented exacerbations of other chronic diseases. For example, patients hospitalized for congestive heart failure and shortness of breath were considered to be symptomatic. Interestingly, the one specific symptom of COVID-19 infection, loss of smell, was not observed in any case. Moreover, protective effectiveness increased over time, which was unexpected. This could be explained by persistent shedding, particularly in those with positive tests close to 90 days after infection. If some reinfections were actually just persistent shedding, then protective efficacy would be higher than what we report.

There are other reasons to believe that immunity will be long-lasting. A study of 705 participants in the United Kingdom with sequential blood sample draws found that 87.8% remained seropositive for at least 6 months after initial infection.^15^ Surprisingly, only 5 participants in that study became negative within 3 months of initial infection. Another study assessing immunological memory in samples from COVID-19 cases found that 95% of subjects had immune memory 6 months after infection, including antibody or T cell responses.^16^ Memory B cells were present for over 6 months in another study.^17^ This suggests that immunity persists beyond the 90-day time period.

Persistent shedding may be characterized by low viral loads or ongoing immune response rather than being a transmissible state. Because asymptomatic transmission is an important means of viral spread, it is crucial to differentiate between asymptomatic reinfection and post-symptomatic viral shedding. If previous infection does not prevent asymptomatic infection, it is possible that previously infected patients could spread the virus, even if they experience no symptoms themselves. Several observational cohorts suggest this is not the case. One study, consisting of mainly non-hospitalized individuals, found that 14.3% of participants were repeatedly positive after recovery. There was no transmission among 757 close contacts of these post-symptomatic carriers.^14^ Another study of 285 patients with positive PCR detected after recovery found that no new cases attributed to those patients occurred in 790 close contacts.^18^

Our study is limited by lack of access to testing results occurring outside of our health system. It is possible that patients were tested for COVID-19 outside the health system, especially if they were asymptomatic, since Cleveland Clinic does not test asymptomatic patients unless they were admitted to hospital or undergoing a procedure/surgery. This would result in an underestimate of the reinfection rate. However, there is no reason to suspect that previously positive patients would be more likely to be retested outside the system than would previously negative patients, which is one reason we included the comparison group. Repeat positive tests could have represented persistent shedding, in which case our estimates of protective effectiveness against true reinfection are too low. Further studies are needed to determine if other SARS-CoV-2 lineages, such as those found in Brazil, South Africa and the United Kingdom, are susceptible to immunity generated from previous infection or vaccination.

In this study of patients in one health system, previous infection appears to offer high levels of protection against symptomatic infection, as well as severe disease, for at least 8 months. In light of these findings, as well as other evidence of the persistence of immunity after infection, the CDC may wish to revisit its recommendation to immediately vaccinate previously infected individuals. Based on this study, patients with known history of infection could delay vaccination for at least 8 months, freeing up vaccine to protect the most vulnerable.

## Data Availability

Further information regarding the dataset is available from the corresponding author upon reasonable request.

